# Walking to the beat: the impact of non-invasive brain stimulation and music on gait in Parkinson’s Disease

**DOI:** 10.64898/2026.04.08.26350408

**Authors:** Marina Emerick, Jessica A. Grahn

## Abstract

Walking impairments in Parkinson’s disease (PD), including reduced speed, cadence, and stride length, along with increased variability, impair mobility and raise fall risk. Conventional treatments may fail to address these deficits, underscoring the need for complementary non-invasive alternatives. This study examined whether combining rhythmic auditory cueing with transcranial direct current stimulation (tDCS) over the supplementary motor area (SMA), a critical region for internally-generated movement, would enhance gait performance in PD. Thirty-three participants with PD and thirty-two healthy controls completed two sessions (anodal vs. sham tDCS) with gait assessed during stimulation, immediately after stimulation, and 15 minutes after stimulation under two auditory conditions: walking in silence and walking to music paced 10% faster than baseline cadence. Spatiotemporal, variability, and stability gait parameters were analyzed using linear mixed-effects models. Rhythmic auditory cueing significantly increased cadence and speed during, immediately after, and especially 15 minutes after stimulation, suggesting sustained effects of rhythmic entrainment. Anodal tDCS produced faster cadence, as well as lower stride time variability and stride width, particularly in individuals with PD. Although both music and anodal tDCS affected gait, no interaction was observed, indicating independent effects. Individuals with PD had greater gait variability overall, and adjusted temporal gait parameters less to music than healthy controls did. Anodal stimulation reduced walking variability in PD, reducing the group differences observed under sham conditions. These findings suggest that rhythmic cueing and SMA stimulation target complementary mechanisms, highlighting the promise of combined tDCS–music interventions for gait rehabilitation in PD.

## Introduction

Walking is essential for daily life, supporting mobility, independence, social interaction, and mental health (Fagerström & Borglin, 2010; Heesch et al., 2015; Patterson & Chapman, 2004). Gait declines with age and neurological disease, increasing falls, hospitalization, and mortality (Ebersbach et al., 2000; Mahlknecht et al., 2013; Ostir et al., 2015; Pirker & Katzenschlager, 2017) . One of the most significant causes of gait impairment among neurological disorders is Parkinson’s disease (PD). Parkinson’s disease is a neurodegenerative disorder characterized by the progressive loss of substantia nigra neurons and reduced dopamine levels in the striatum, a brain region crucial for motor control (Björklund & Dunnett, 2007; Groenewegen, 2003). These neurobiological changes lead to motor symptoms such as bradykinesia, postural instability, muscle rigidity, and resting tremor (Hughes et al., 1992), as well as nonmotor symptoms such as depression, sleep disturbances, and fatigue (Salawu et al., 2010).

Gait disturbances are among the most disabling PD symptoms, contributing to reduced mobility and greater fall risk (Ashoori et al., 2015; Gazibara et al., 2015). Compared to healthy adults, individuals with PD typically show reduced walking speed, cadence (steps per minute), and stride length, as well as higher stride variability (Blin et al., 1990; Morris et al., 1994, 1996). In later stages, freezing of gait (FOG)—a transient inability to initiate or maintain movement despite the intention to walk—often emerges (Grabli et al., 2012). FOG is associated with poorer cognitive functioning and reduced quality of life (Perez-Lloret et al., 2014), and has been linked to disruptions in internal rhythmic timing networks (Tolleson et al., 2015).

PD treatments include pharmacotherapy such as Levodopa, a dopaminergic precursor, and deep brain stimulation (DBS) to improve the patient’s overall motor symptoms. However, these treatments have a limited effect on gait impairments (Bella et al., 2015; Blin et al., 1990; Grabli et al., 2012). Moreover, DBS is an invasive procedure with surgical risks, and pharmacological treatments may induce side effects such as drowsiness and dyskinesias that can worsen gait deficits (Jankovic, 2008, 2015; Nombela et al., 2013). Consequently, alternative methods for improving gait disturbances have been proposed to re-establish connections within these disrupted internal rhythmic networks, such as rhythmic auditory cueing (Thaut, 2005). By providing auditory cues such as metronomes or music with a clear beat, rhythmic cueing helps individuals synchronize their movements to the external rhythm, improving stride velocity, stride length, and overall gait pattern (Thaut et al., 2015). Evidence supports the effectiveness across various healthy and clinical populations (Arias & Cudeiro, 2010; Ghai et al., 2018; Lim et al., 2005; Ready et al., 2022; Rochester et al., 2005, 2010; Thaut & Abiru, 2010).

Gait improvements with rhythmic cueing may arise from interactions between auditory and motor systems, particularly two key pathways: the basal ganglia–supplementary motor area (SMA) circuit and the cerebellar–thalamo–cortical circuit (Nombela et al., 2013). The basal ganglia–SMA loop normally supports internally generated timing and automatic sequencing of movements (Cannon & Patel, 2021; Morris et al., 1996). In Parkinson’s disease, dysfunction of the basal ganglia reduces SMA activity (Jahanshahi et al., 1996; Sabatini et al., 1998), leading to impaired self-paced movement initiation and timing (Muthukrishnan et al., 2019). When this internal timing mechanism is compromised in PD, movement becomes less automatic and more variable. Rhythmic auditory cues may compensate by providing an external temporal template, allowing individuals to synchronize steps with an external beat instead of relying solely on impaired internal timing systems (Morris et al., 1996; Muthukrishnan et al., 2019). Neuroimaging evidence shows that externally cued movements recruit premotor and cerebellar regions (Hallett, 2008; Taniwaki et al., 2006) potentially bypassing or complementing the impaired basal ganglia–SMA network (Nombela et al., 2013). Thus, rhythmic music may support gait by engaging cerebellar–thalamo–cortical pathways and reducing reliance on deficient automatic control circuits in PD.

A useful method to investigate the neural mechanisms of auditory cueing effects on gait in PD is transcranial direct current stimulation (tDCS), a non-invasive technique that passes a low electrical current between two scalp electrodes to modulate cortical excitability. Depending on the direction of current flow, excitability in underlying brain tissue can be increased or decreased (Poreisz et al., 2007; Reinhart et al., 2017). Because it transiently alters neural activity, tDCS can provide causal evidence of the role of a specific brain area in a particular function. It has been used in gait studies of motor adaptation, dual-task walking, force production, somatosensory function, and walking mobility/coordination with younger adults (Fernandez et al., 2017; Koganemaru et al., 2018; Orcioli-Silva et al., 2021; Van Asseldonk & Boonstra, 2016; Wrightson et al., 2015; Zhou et al., 2014), older adults (Orcioli-Silva et al., 2021; Rostami et al., 2020; Schneider et al., 2021; Zhou et al., 2018), and individuals with PD (Pol et al., 2021). Repeated tDCS sessions produce stronger and more sustained effects when paired with tasks that engage the same brain regions targeted by stimulation (Berryhill & Martin, 2018; Brasil-Neto, 2012; Jones et al., 2017; Park et al., 2014; Schneider et al., 2021; Stephens & Berryhill, 2016, 2016). In gait research, multi-session tDCS protocols combined with training yield greater improvements than single-session interventions in healthy older adults (Rostami et al., 2020).

The timing of stimulation relative to task performance—whether tDCS is applied during (online) or before/after (offline) gait training—may also influence the immediacy and magnitude of its effects. Evidence suggests that applying tDCS during gait- or dual-task training can facilitate activity-dependent plasticity and reduce dual-task costs (Koganemaru et al., 2018; Mitsutake et al., 2021; Schneider et al., 2021). In contrast, some studies report improvements emerging after stimulation, with peak effects observed around 15 minutes post-stimulation (Mishra & Thrasher, 2021). Others have found that stimulation applied before walking may prime the motor system, although results remain mixed depending on task demands (Workman et al., 2019). These findings indicate that both online and offline tDCS may modulate gait outcomes; however, evidence remains inconsistent, and further work is needed to characterize their respective effects.

Building on this evidence, the present study aims to address a key gap in the literature: while both tDCS and rhythmic auditory cueing have independently shown promise in improving gait, their combined effects remain unclear, particularly in PD. To explore this, we paired anodal tDCS over the SMA with rhythmic auditory cueing, using music containing a clear beat, during gait. This design was chosen to target both internal and external timing mechanisms of gait control. Specifically, anodal stimulation of the SMA was expected to enhance internally generated movement initiation and sequencing, which are impaired in PD (Jahanshahi et al., 1996; Sabatini et al., 1998), while rhythmic auditory cues were hypothesized to support externally guided movement through cerebellar–thalamo–cortical pathways (Nombela et al., 2013). Thus, SMA activation may complement auditory cueing by strengthening top-down motor timing or interact with it by amplifying the effects of rhythmic entrainment on gait.

To examine these possibilities, participants completed two sessions—one with active anodal tDCS and one with sham stimulation—while walking to rhythmic music. We assessed gait during stimulation, immediately after, and 15 minutes post-stimulation to capture potential differences in the time course of effects. We hypothesized that (1) if anodal tDCS over the SMA enhances the effect of auditory cues, then combining tDCS with rhythmic music would enhance gait velocity, cadence, and stability and reduce variability beyond the additive effects of each intervention alone, indicating an interaction effect between brain stimulation and auditory cueing. Alternatively, (2) if tDCS and rhythmic cueing act independently to improve gait, we should observe additive main effects of each intervention—both will improve gait velocity, cadence, and step-to-beat matching, but their combination will not be super-additive.

This study will also provide insights into the role of the SMA in mediating the effects of RAS on gait, deepening our understanding of how music-driven auditory-motor interactions can enhance movement in PD. Understanding the mechanisms by which auditory cues influence movement is crucial, as gait disturbances significantly impact the quality of life for PD individuals. The findings could inform the development of innovative treatments, such as combining non-invasive brain stimulation with music rehabilitation programs, to address motor impairments in PD.

## Methods

### Participants

We recruited 36 participants diagnosed with Parkinson’s Disease. Three participants were excluded due to non-compliance with task instructions, atypical baseline performance, or preprocessing error, resulting in a final sample of 33. We also recruited 33 older adults without PD. One was excluded for non-compliance with task instructions, resulting in a final sample of 32. Demographic data for both groups are shown on Table 1.

**Table 1.** Participants’ demographic data.

|  | PD Group (N = 33) | Control Group (N = 32) |
| --- | --- | --- |
| Age (years) | 67 (± 10.8) | 67.3 (± 7.7) |
| Sex (male/female) | 17/16 | 11/21 |
| Goldsmith Musical Training Scale | 17.1 (± 11.5) | 14.9 (± 9.9) |
| Dance Skills (%) | 90.9% no<br>experience/beginner<br>9.1% intermediate | 81.2 % no<br>experience/beginner<br>15.6 % intermediate<br>3.2 % advanced |
| Montreal Cognitive Assessment 8.1 | 27 (± 2.4) | 26.8 (± 2.2) |
| H&Y Score (mean ± SD) | 1.3 (± 1) | NA |
| MDS-UPDRS II Score (mean ± SD) | 12.3 (± 6.3) | NA |
| MDS-UPDRS III Score (mean ± SD) | 23.6 (± 13) | NA |
| Disease duration (years ± SD) | 4.8 (± 3.1) | NA |
| Timed Up-and-Go Test (mean ± SD) | 11.6 (± 3.3) | NA |

Participants were recruited through the Parkinson Society Southwestern Ontario, Western University’s OurBrainsCAN Registry, and by word of mouth. The study was approved by the Health Sciences Research Ethics Board at Western University, and conducted in accordance with relevant guidelines (Bikson et al., 2009; Nitsche et al., 2008).

To be included in the study, participants were required to have a confirmed diagnosis of Parkinson’s disease (for the Parkinson’s group), no history of serious brain trauma, and sufficient physical, visual, and cognitive ability to complete the study (Dobbs et al., 2018). Exclusion criteria were contraindications to tDCS, including a history of epilepsy, metal implants in the head or neck, skin conditions near the stimulation site, or previous adverse reactions to brain stimulation. (Pilloni et al., 2022; Thair et al., 2017).

All individuals with PD were in the early stages of the disease (Hoehn & Yahr stages 1–3) and were able to walk unassisted. To ensure consistent functional capacity, PD participants were tested in the ON state of their medication.

## Materials

### Questionnaires

A demographic questionnaire assessed age, educational level, language, general health, and musical and dance abilities, as well as PD information, such as diagnosis period, medication type and schedule, and the perceived impact of PD on participants’ daily lives.

Musical training was assessed with the musical training subscale from the Goldsmiths Musical Sophistication Index (Müllensiefen et al., 2014), which has seven items, with scores ranging from 7 to 49.

Participants listened to 30-s clips of the eight musical stimuli used for the walk trials and rated them on familiarity, groove, enjoyment, and beat salience on 100-point Likert scales.

Cognitive function was evaluated using the Montreal Cognitive Assessment (MoCA) (Nasreddine et al., 2005). Motor abilities and disease severity were assessed using parts II and III of the Unified Parkinson’s Disease Rating Scale (UPDRS) (Goetz et al., 2008). Disease stage was classified according to the Hoehn and Yahr (H&Y) scale (Hoehn & Yahr, 1967). Mobility was measured with the Timed Up & Go (TUG) test (Podsiadlo, 1991) in participants with PD only.

### Beat Alignment Test - BAT

Beat perception and production abilities were assessed using the Beat Alignment Test (BAT) (Müllensiefen et al., 2014). In the perception task, participants listened to short excerpts of music with a metronome-like sequence of beeps overlaid and indicated whether the beeps matched the underlying beat of the music or not. Performance was quantified as the percentage of correct responses.

In the production task, participants tapped in time with the beat of various musical excerpts. We extracted measures of asynchrony (the temporal deviation between the participant’s taps and the musical beat) and the coefficient of variation (CoV) of the participant’s inter-tap intervals. All excerpts were presented in randomized order, and participants were instructed to make their judgments without moving their bodies to the rhythm.

At the end of each session, participants completed a short questionnaire assessing their experience with tDCS sensations and their perceived ability to synchronize their steps to the beat. Specifically, participants were asked: (1) how difficult it was to synchronize their steps to the beat, rated on a scale from 0 (least difficult) to 100 (most difficult); (2) whether they noticed any unusual sensations under the stimulation electrodes (Sensation); and if yes (3) the intensity of any such sensations, rated on a 0 (least sensation) to 100 (most sensation) scale.

### Auditory Stimuli

Auditory stimuli were delivered via Sennheiser RS 175 Bluetooth headphones and consisted of eight non-lyrical, familiar, high-groove songs (Table 2). Each song was edited to remove lyrics and tempo-adjusted in Audacity (Audacity Team, 2024, v3.5.1), with pitch preserved, to be 10% faster than the participant’s baseline cadence (e.g., if the baseline was 100 steps per minute, the song was adjusted to 110 beats per minute).

**Table 2.**
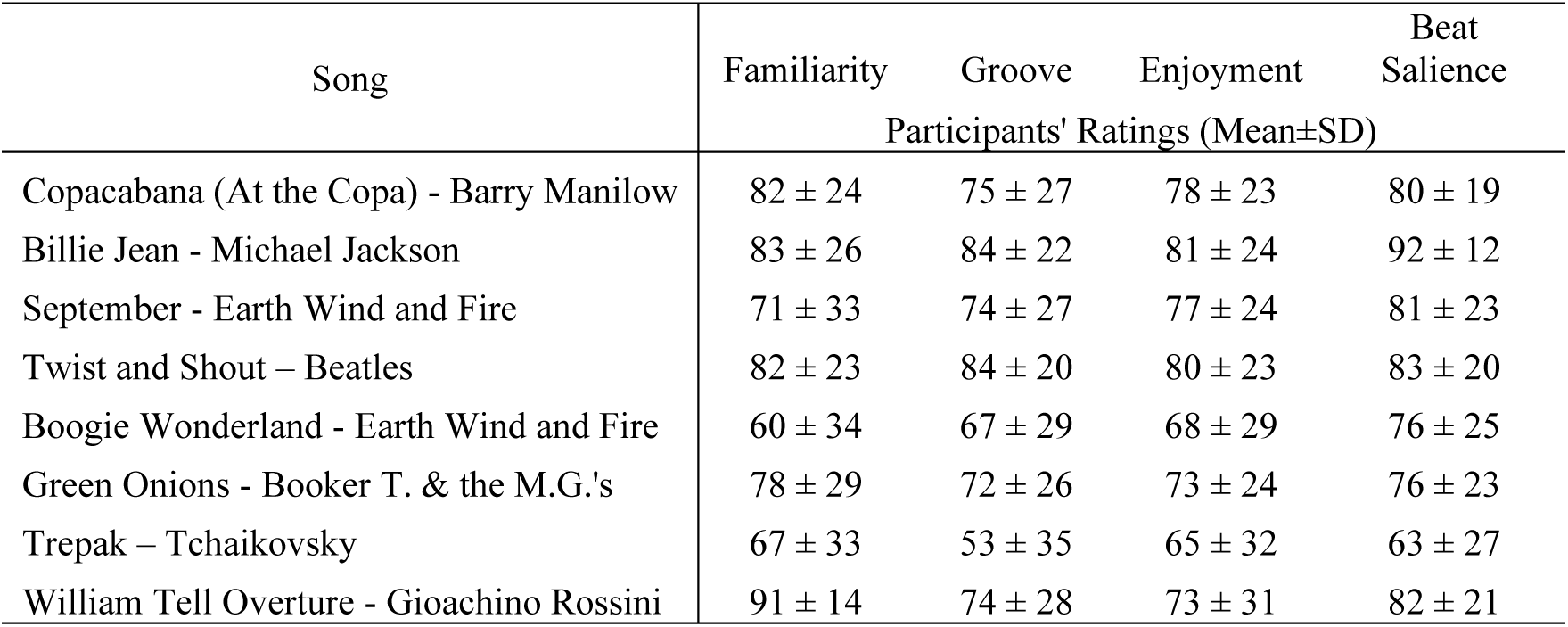
Music cueing stimuli and participants’ ratings of familiarity, groove, enjoyment and beat salience.

### Transcranial Direct Current Stimulation (tDCS)

Non-invasive brain stimulation was delivered using an Activadose system (Caputron). Each electrode (4.5 × 4.5 cm) was inserted into a 5 × 5 cm rubber cap containing a saline-soaked sponge (0.9% NaCl; effective contact area = 3 × 3 cm). A 2-mA current was applied for 20 minutes, resulting in a current density of 0.22 mA/cm². Electrodes were secured with rubber head straps. For the active tDCS condition, the current was gradually ramped up to 2 mA over 30 s at the start of the task, maintained for 20 min, and then ramped down. For the sham condition, stimulation was similarly ramped up over 30 s to 2 mA but immediately ramped back down to 0 mA over the next 30 s. This procedure mimics the initial sensation of stimulation without producing lasting neuromodulatory effects (Ambrus et al., 2012). Electrodes were positioned according to the international 10–20 system and following the procedures described by Emerick et al. (2025). The anodal electrode was placed over SMA, located 2 cm anterior to Cz (at the vertex of the scalp), and the reference electrode was positioned on the forehead above the right eye.

### Gait measurement

Gait data were acquired using a 4.9-meter pressure-sensitive walkway (Zeno Walkway, ProtoKinetics). For each walking trial, participants walked continuously in a loop, stepping an additional 1.78 m beyond each end of the mat before turning around, as described by Ready et al. (2025). Each trial consisted of six passes (three back-and-forth walks) across the walkway. In the silent walks, participants were instructed to walk at their normal pace, beginning and ending each trial upon the researcher’s verbal cue. During the music walks, participants were instructed to synchronize their steps with the beat of the music played through headphones, and began walking once they perceived the beat in the music, and stopped when the music ended.

### Procedure

Participants received sham and anodal stimulation in two counterbalanced sessions on different days. In each session, baseline gait was measured following the same procedures as in Ready et al. (2022); participants completed one walking trial on the gait mat in silence at a comfortable, self-selected pace. Then, they completed the song ratings questionnaire if on day 1, and the demographic questionnaire if on day 2. Afterward, they completed 6 silent walking trials and 6 walking trials with music (6 songs) that was matched to 10% faster than their baseline cadence. The walking and music conditions each took 10 minutes, during which tDCS (either sham or anodal) was applied. Short breaks were given between each silent and music walk. They also completed one silent trial and one music trial immediately after the tDCS-stimulated trials, and again 15 minutes after tDCS. Thus, participants completed trials under the following conditions: baseline, silent walking/sham tDCS, music walking/sham tDCS, silent walking/anodal tDCS, and music walking/anodal tDCS. During the 15-minute break between the post-stimulation walks and the final walks, participants completed the beat alignment task (BAT) and the TUG test. After the last 2 walks, participants completed the MoCA test on day 1, and the UPDRS parts II and III on day 2.

## Data analysis

For measures other than gait variability, a normalized score was calculated to account for individual differences. The proportional change from baseline was calculated for cadence, stride length, stride velocity, stride width and double support time.

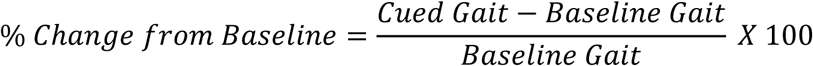

Parameters were calculated for each condition during, after and 15-minutes after tDCS: silent walking/sham tDCS, music walking/sham tDCS, silent walking/anodal tDCS, and music walking/anodal tDCS.

### Statistical Analysis

Statistical analyses were conducted in R (version 4.3.2) using the lme4, lmerTest, emmeans, and performance packages (Bates et al., 2015; Kuznetsova et al., 2017; Lenth, 2016; Lüdecke et al., 2021). Because each participant contributed repeated measurements across the different walking conditions, all gait outcomes were analyzed using linear mixed-effects models (LMMs), with participant ID entered as a random intercept to account for within-subject dependence.

For each gait parameter (cadence, stride length, stride velocity, stride width, double support time, and the coefficients of variation for stride time and stride length), a model was estimated including the fixed effects of stimulation type (anodal, sham), cue (music, silent), time (during stimulation, post-stimulation, 15-min post-stimulation), and group (healthy older adults - OA, Parkinson’s disease - PD), as well as their interactions.

For individual difference factors, dimensionality reduction was performed prior to modeling. Principal component analysis (PCA) was conducted on rhythm and music-related measures: beat perception accuracy, beat production asynchrony, beat production coefficient of variation, and Goldsmith Musicality scores. We used the first principal component (PC1) in the model (Rhythm). Higher PC1 scores reflected better sensorimotor rhythmic performance, characterized primarily by lower tapping asynchrony, lower tapping variability, and higher musicality scores. Rhythm was entered as a fixed effect to test whether rhythmic abilities moderated responsiveness to rhythmic cueing, brain stimulation, time, and group interactions. Age and sex were included as covariates. All continuous predictors were mean centered prior to analysis. The model was specified as:

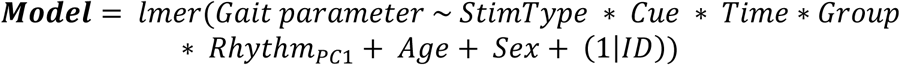

All models were fit using maximum likelihood estimation to allow valid comparisons across nested model structures.

Fixed effects in all mixed-effects models were evaluated using Type III ANOVA with Satterthwaite–Kenward–Roger degrees of freedom, as implemented in the *lmerTest* package. Statistical significance was set at p < .05 (two-tailed).

When significant main effects or interactions were detected, follow-up comparisons were conducted using estimated marginal means computed with the *emmeans* package. Pairwise comparisons between levels of categorical predictors (stimulation type, cue, time, and group) were performed with Tukey-adjusted p-values to control for multiple comparisons. For interactions involving continuous moderators, such as Rhythm, simple slope analyses were conducted using estimated marginal trends (*emtrends*) to evaluate whether the relationship between the moderator and the gait outcome differed across different levels.

Effect sizes appropriate for mixed models were reported using semi-partial 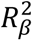 values computed for each fixed effect using *r2beta* function (method = "kr"), which applies the Kenward–Roger approximation.

### Parkinson’s Disease – Specific Analysis

To further examine the role of disease severity, a secondary set of models was conducted including only participants with PD. A separate PCA was performed on UPDRS motor scores, Hoehn and Yahr stage, and disease duration, and the first principal component (Disease PC1) was retained as an index of overall disease severity. Disease severity component was included as a continuous predictor, both as a main effect and in interaction with stimulation type and cue, to test whether disease severity moderated responsiveness to auditory cueing and stimulation type. Model structure and statistical procedures were otherwise identical to those described for the primary analysis, and significant results involving disease severity were reported for each gait parameter.

## Results

### Spatiotemporal variables Cadence

For cadence, there were significant main effects of stimulation, cue, and time. Cadence was higher during anodal than sham stimulation (F(1,712) = 19.05, p < .001, R²β = .026), and higher during music than silence (F(1,712) = 553, p < .001, R²β = .437). Cadence also varied across time (F(2,712) = 24.43, p < .001, R²β = .064), with the highest value at 15 minutes post-stimulation compared to both during stimulation (p < .0001) and immediately post-stimulation (p < .0001), while the latter two did not differ (p = .529). Participants with stronger rhythmic abilities showed greater cadence overall (F(1,65) = 5.02, p = .029, R²β = .072). There was no significant main effect of group (F(1,65) = 1.58, p = .213, R²β = .024). (Figure 1.A)

**Figure 1.**
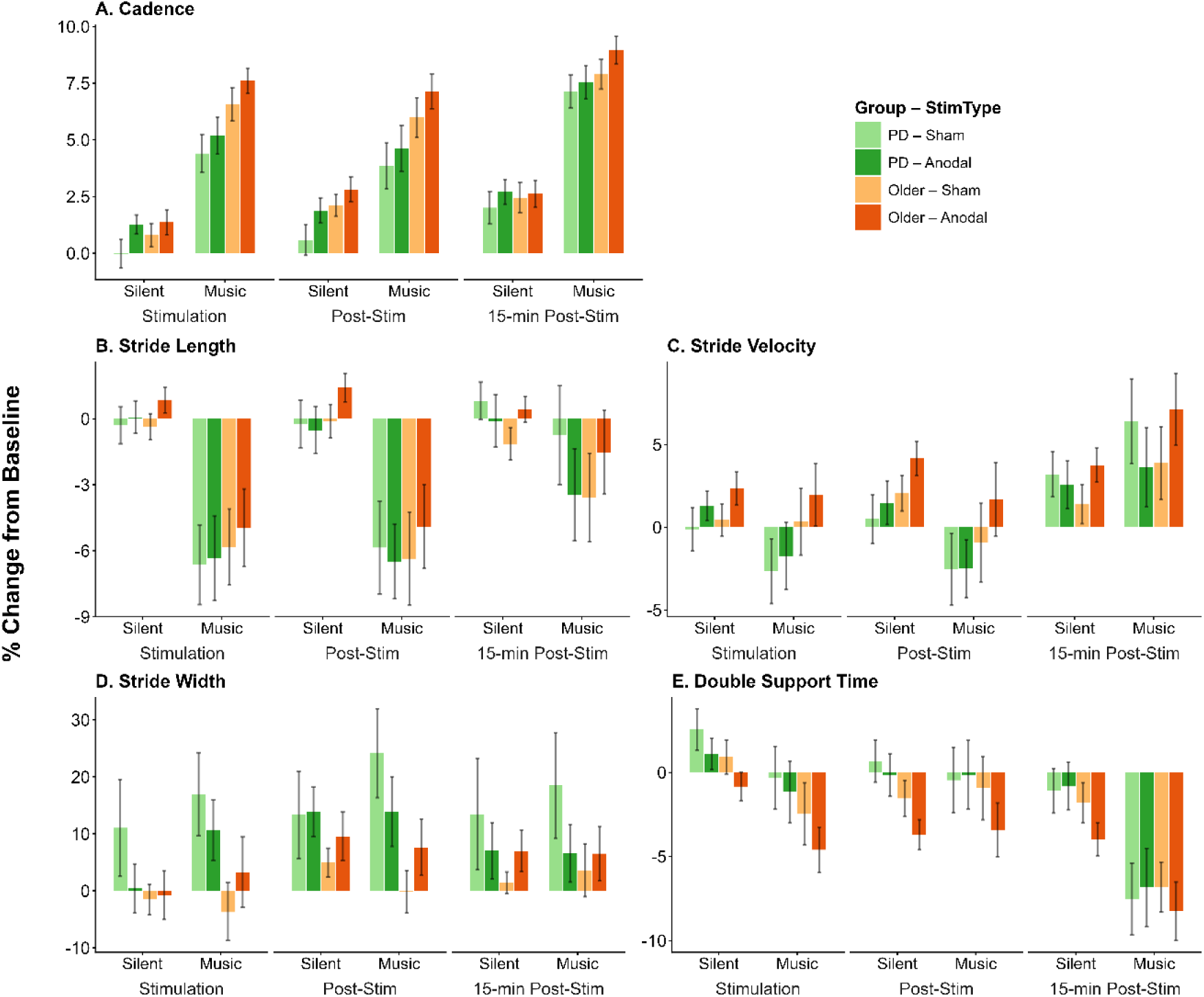
Spatiotemporal and stability gait variables in individuals with Parkinson’s disease and healthy older adults. Mean percent change from baseline for (A) cadence, (B) stride length, (C) stride velocity, (D) stride width, and (E) double support time during silent and music walking conditions across three time points: during stimulation, post-stimulation, and 15-min post-stimulation. Bars represent two groups—individuals with PD (green hues) and older adults (orange hues)—under two stimulation conditions: sham (lighter shades) and anodal (darker shades). Error bars indicate ±1 standard error of the mean (SEM).

**Figure 2.**
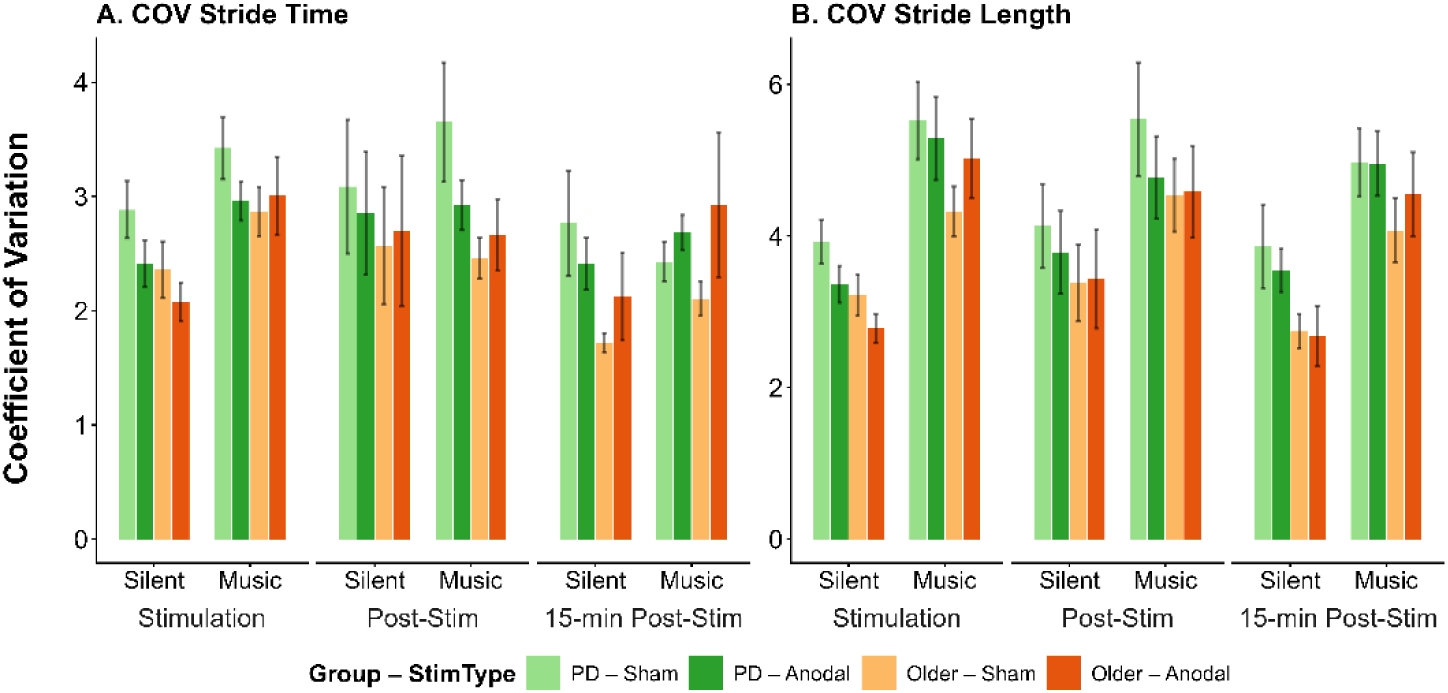
Variability of gait variables in individuals with Parkinson’s disease and healthy older adults. Changes in coefficient of variation (COV) for (A) stride time and (B) stride length, during silent and music walking conditions across three time points: during stimulation, post-stimulation, and 15-min post-stimulation. Bars represent two groups—individuals with PD (green hues) and older adults (orange hues)—under two stimulation conditions: sham (lighter shades) and anodal (darker shades). Error bars indicate ±1 standard error of the mean (SEM).

These effects were qualified by a significant Cue × Time interaction (F(2,712) = 6.81, p = .001, R²β = .02), in which the difference between music and silence was lower during stimulation relative to immediately post-stimulation (p = .004) , and higher at 15 minutes post-stimulation relative to immediately post-stimulation (p = .001). There was also a Cue × Group interaction (F(1,712) = 8.58, p = .004, R²β = .01), in which the music effect was smaller in the PD group (p = .005), with post-hoc analysis showing that healthy older adults walked with faster cadence than the PD group during music (p = .04), but not during silence (p = .82). A significant Group × Rhythm interaction (F(1,65) = 4.99, p = .03, R²β = .07) indicated that the relationship between cadence and rhythmic ability differed between groups; post-hoc analysis showed a near zero trend in older adults (slope = 0.00, SE = 0.38, 95% CI [−0.76, 0.76]), but a significant positive relationship in the PD group (slope = 1.15, SE = 0.38, 95% CI [0.39, 1.91]), with the difference between groups reaching significance (Older − PD = −1.15, SE = 0.54, p = .04). This was further qualified by a significant Cue × Group × Rhythm interaction (F(1,712) = 26.07, p < .001, R²β = .04). In older adults, the association between cadence and rhythmic ability was positive during silence (slope = 0.39, SE = 0.40, 95% CI [−0.40, 1.18]) but negative during music (slope = −0.39, SE = 0.40, 95% CI [−1.18, 0.41]); the difference between conditions was significant (Silent Older − Music Older = 0.78, SE = 0.25, p = .01), indicating that music reversed the direction of the rhythmic ability and cadence relationship in older adults. In the PD group, the pattern was the opposite: higher rhythmic ability predicted faster cadence during music (slope = 1.63, SE = 0.40, 95% CI [0.83, 2.42]) more strongly than during silence (slope = 0.67, SE = 0.40, 95% CI [−0.13, 1.46]). The difference between conditions was significant (Silent PD − Music PD = −0.96, SE = 0.25, p = .001), indicating that music accentuated the rhythmic ability effect on increasing cadence in PD. The music rhythm slope in PD was significantly steeper than the music rhythm slope in older adults (Music PD vs. Music Older: p = .003). (Figure 3.A)

**Figure 3.**
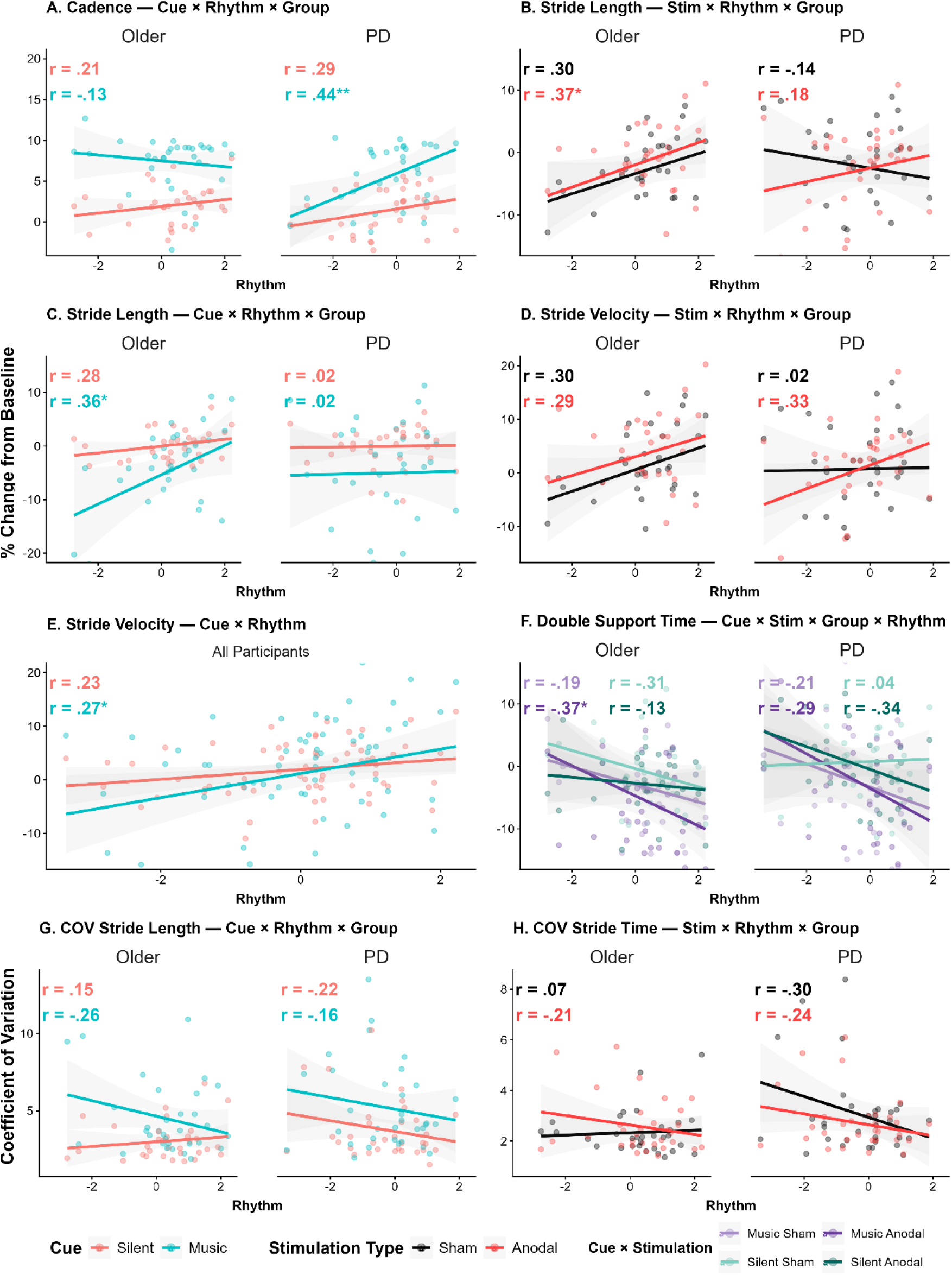
Effects of auditory cueing and stimulation on gait outcomes as a function of rhythmic ability across healthy older adults and individuals with Parkinson’s disease. Scatterplots display relationships between rhythm score (x-axis) and gait outcomes (y-axis), expressed as percent change from baseline or coefficient of variation (COV), depending on the panel. Solid lines indicate linear regressions with shaded 95% confidence intervals; correlation coefficients (R) and p-values are shown within each panel. (A) Cadence: Cue × Rhythm × Group interaction, shown separately for older adults and PD. (B) Stride length: Stimulation × Rhythm × Group interaction, shown separately for older adults and PD. (C) Stride length: Cue × Rhythm × Group interaction, shown separately for older adults and PD. (D) Stride velocity: Stimulation × Rhythm × Group interaction, shown separately for older adults and PD. (E) Stride velocity: Cue × Rhythm interaction across groups. (F) Double support time: Cue × Stimulation × Group × Rhythm interaction. (G) COV stride length: Cue × Rhythm × Group interaction. (H) COV stride time: Stimulation × Rhythm × Group interaction. Color coding reflects experimental conditions: cue type (silent vs. music; teal vs. peach) and/or stimulation type (sham vs. anodal; black vs. red), with combined cue × stimulation conditions indicated in panel F.

### Stride Length

For stride length, there were significant main effects of cue, time, and sex. Stride length was shorter during music walking (F(1,712) = 132.00, p < .001, R²β = .16) than silence. In terms of time (F(2,712) = 5.69, p = .004, R²β = .02), strides were longer at 15 minutes post-stimulation compared to both during stimulation (p = .013) and immediately post-stimulation (p = .014), with no difference between the latter two (p = .999). The change in stride length from baseline differed by sex, where females showed longer strides than males (F(1,65) = 5.58, p = .02, R²β = .08). There were no significant main effects of stimulation type (F(1,712) = 2.37, p = .12), group (F(1,65) = 0.28, p = .60), or rhythm (F(1,64) = 2.32, p = .13). (Figure 1.B) These effects were qualified by a significant Cue × Time interaction (F(2,712) = 7.06, p < .001, R²β = .02), in which the reduction in stride length during music walking relative to silence was larger at 15 minutes post-stimulation than during stimulation (p = .002) and immediately post-stimulation (p = .002), with no difference between the latter two (p = .98). Significant interactions were observed for Stimulation × Rhythm (F(1,712) = 8.92, p = .003, R²β = .01) and Cue × Rhythm (F(1,712) = 8.88, p = .003, R²β = .01), but are not interpreted in light of significant three-way interactions. A significant Stimulation × Group × Rhythm interaction (F(1,712) = 6.24, p = .01, R²β = .01) was present. Post-hoc analysis showed that in the PD group, the rhythm slope was significantly more positive during anodal stimulation (slope = 1.13, SE = 0.89, 95% CI [−0.64, 2.90]) than during sham (slope = −0.85, SE = 0.89, 95% CI [−2.62, 0.92]).

The difference between conditions was significant (Sham PD − Anodal PD = −1.98, SE = 0.53, p = .001), indicating that anodal stimulation amplified the rhythmic ability benefit for stride length in PD. No such difference was observed in older adults (Sham Older slope = 1.51, SE = 0.89, 95% CI [−0.25, 3.27] vs. Anodal Older slope = 1.69, SE = 0.89, 95% CI [−0.08, 3.45]; Sham Older − Anodal Older = −0.18, SE = 0.53, p = .99) (Figure 3.B). A Cue × Group × Rhythm (F(1,712) = 8.20, p = .004, R²β = .01) interaction showed that among older adults, the rhythm slope was significantly more positive during music than silence; higher rhythmic ability predicted longer strides during music walking (slope = 2.65, SE = 0.89, 95% CI [0.89, 4.41]), whereas this relationship was much weaker during silent walking (slope = 0.55, SE = 0.89, 95% CI [−1.22, 2.31]). The difference between conditions was significant (Silent Older − Music Older = −2.10, SE = 0.53, p < .001), indicating that music accentuated the rhythmic ability benefit for stride length in older adults. In the PD group, rhythmic ability was not differentially associated with stride length across cue conditions; the rhythm slope was similarly minimal during silent (slope = 0.12, SE = 0.89, 95% CI [−1.65, 1.89]) and music walking (slope = 0.16, SE = 0.89, 95% CI [−1.61, 1.93]), and the difference was non-significant (Silent PD − Music PD = −0.04, SE = 0.53, p = 1.00). (Figure 3.C).

Additionally, the PD specific analysis showed a significant interaction between cue and disease severity (F(1,362) = 12.56, p < .001, R²β = .03). Post-hoc analysis showed that higher disease severity predicted a steeper reduction in stride length during music walking (slope = −1.69, SE = 0.95, 95% CI [−3.60, 0.21]), whereas this relationship was minimal during silent walking (slope = −0.11, SE = 0.95, 95% CI [−2.02, 1.79]). The difference between conditions was significant (Silent − Music = 1.58, SE = 0.46, p < .001), indicating that music accentuated disease-related reductions in stride length (Figure 4.A).

**Figure 4.**
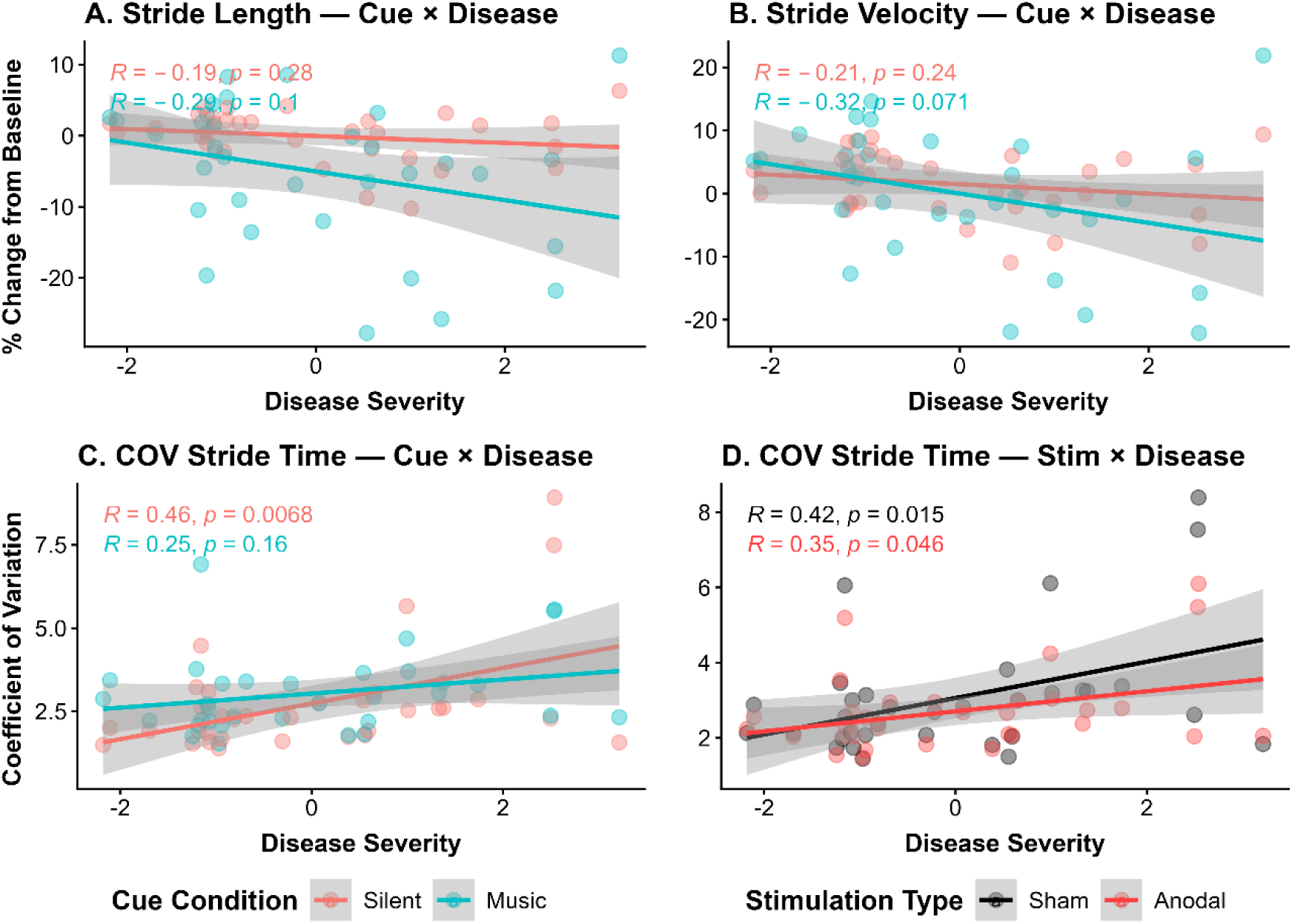
Effects of auditory cueing and stimulation on gait outcomes as a function of disease severity in individuals with Parkinson’s disease. Scatterplots display relationships between disease severity (x-axis) and gait outcomes (y-axis), expressed as percent change from baseline or coefficient of variation (COV), depending on the panel. Solid lines indicate linear regressions with shaded 95% confidence intervals; correlation coefficients (R) and p-values are shown within each panel. (A) Stride length: Cue × Disease interaction. (B) Stride velocity: Cue × Disease interaction. (C) COV stride time: Cue × Disease interaction. (D) COV stride time: Stimulation × Disease interaction. Color coding reflects experimental conditions: cue type (silent vs. music; teal vs. peach) and stimulation type (sham vs. anodal; black vs. red), as indicated in each panel.

### Stride Velocity

For stride velocity, there were significant main effects of stimulation, time, rhythm, and sex. Velocity was higher during anodal than sham stimulation (F(1,712) = 8.94, p = .003, R²β = .012), and higher at 15 minutes post-stimulation compared to both during stimulation (p < .001) and immediately post-stimulation (p < .001), which did not differ (p = .909; F(2,712) = 19.27, p < .001, R²β = .051). Participants with higher rhythmic abilities walked faster overall (F(1,65) = 5.02, p = .029, R²β = .072), and females walked faster than males (F(1,65) = 7.09, p = .010, R²β = .098; post-hoc: p = .013). There was no significant main effect of cue (F(1,712) = 2.55, p = .110) or group (F(1,65) = 0.001, p = .979). (Figure 1.C)

These effects were qualified by a significant Cue × Time interaction (F(2,712) = 9.95, p < .001, R²β = .027), in which the music-silence difference in velocity was larger at 15 minutes post-stimulation compared to during stimulation (p = .003) and immediately post-stimulation (p < .001), with no difference between the latter two (p = .213). A significant Cue × Rhythm interaction (F(1,712) = 9.37, p = .002, R²β = .013) indicated that the positive relationship between rhythmic ability and velocity was stronger during music than silence. Post-hoc analysis showed that higher rhythmic ability predicted faster velocity during music walking (slope = 2.124, SE = 0.733, 95% CI [0.668, 3.580]), whereas this relationship was weaker during silent walking (slope = 0.869, SE = 0.733, 95% CI [−0.588, 2.326]). The difference between conditions was significant (Silent − Music = −1.255, SE = 0.424, p = .003), indicating that music accentuated the rhythmic ability effect on walking speed (Figure 3.D). Significant Stimulation × Rhythm (F(1,712) = 4.90, p = .027, R²β = .007), and Stimulation × Group × Rhythm (F(1,712) = 8.31, p = .004, R²β = .012) interactions were observed. Post-hoc contrasts showed that in the PD group, the rhythm slope was significantly more positive during anodal stimulation (slope = 2.278, SE = 1.040, 95% CI [0.212, 4.344]) than during sham (slope = 0.187, SE = 1.040, 95% CI [−1.879, 2.253]). The difference between conditions was significant (Sham PD − Anodal PD = −2.091, SE = 0.598, p = .003), indicating that anodal stimulation amplified the rhythmic ability benefit for walking velocity in PD. No such difference was observed in older adults (Sham Older slope = 1.898, SE = 1.030, 95% CI [−0.159, 3.955] vs. Anodal Older slope = 1.624, SE = 1.030, 95% CI [−0.434, 3.682]; Sham Older − Anodal Older = 0.275, SE = 0.600, p = .968). All other group and stimulation interactions were non-significant (all ps > .190). (Figure 3.E).

Additionally, the PD only model showed a significant interaction between cue and disease severity (F(1,362) = 9.77, p = .002, R²β = .03). Greater disease severity was associated with larger decreases in stride velocity during music walking (slope = −1.61, SE = 1.01, 95% CI [−3.64, 0.42]), relative to silent walking (slope = −0.006, SE = 1.01, 95% CI [−2.03, 2.02]). The contrast between slopes was significant (p = .0024) (Figure 4.B).

### Variability variables Stride Time Variability

For stride time variability (CoV), there were significant main effects of cue and time. Variability was higher during music than silence (F(1,712) = 8.82, p = .003, R²β = .01), and varied across timepoints (F(2,712) = 3.44, p = .03, R²β = .01), with post-stimulation values being higher than 15-minute post-stimulation (p = .05), while the stimulation and post-stim timepoints did not differ (p = .90). (Figure 2.A). A significant Time × Rhythm interaction (F(2,712) = 3.14, p = .04, R²β = .01) showed that the negative relationship between rhythmic ability and stride time variability was most pronounced at 15 minutes post-stimulation (slope = −0.39, SE = 0.14, 95% CI [−0.66, −0.12]) relative to during stimulation (slope = −0.10, SE = 0.14) and immediately post-stimulation (slope = −0.12, SE = 0.14), though pairwise comparisons among slopes did not reach significance (all ps > .08). A significant Stimulation × Group interaction (F(1,712) = 5.16, p = .02, R²β = .01) indicated that anodal stimulation differentially affected stride time variability across groups, decreasing it for the PD group only. The Stimulation × Group × Rhythm (F(1,712) = 4.08, p = .04, R²β = .01) interaction indicated that in the PD group, higher rhythmic ability was associated with lower variability under sham stimulation (slope = −0.44, SE = 0.18, 95% CI [−0.78, −0.09]) than anodal stimulation (slope = −0.24, SE = 0.18, 95% CI [−0.59, 0.11]). The difference between conditions was non-significant (Sham PD − Anodal PD = −0.20, SE = 0.15, p = .58), though the pattern suggests anodal tDCS attenuated the rhythmic ability benefit for stride time variability in PD. In older adults, slopes were near zero for both conditions (Sham Older slope = 0.05, SE = 0.18, 95% CI [−0.30, 0.40]; Anodal Older slope = −0.18, SE = 0.18, 95% CI [−0.53, 0.17]), and no pairwise contrast reached significance after correction (all p > .21) (Figure 3.G).

Additionally, greater disease severity (DiseasePC1) was associated with greater stride time variability (F(1,32.87) = 5.84, p = .021, R²β = .151). Stimulation type interacted with disease severity (F(1,361.9) = 4.19, p = .041, R²β = .011), such that the relationship between disease severity and variability was weaker during anodal (slope = 0.29, SE = 0.19, 95% CI [−0.09, 0.66]) than sham stimulation (slope = 0.51, SE = 0.19, 95% CI [0.13, 0.88]). The difference was significant (Sham − Anodal = 0.22, SE = 0.11, p = .046), (Figure 4. D). Cue and disease severity also interacted (F(1,361.90) = 8.77, p = .003, R²β = .024), such that the relationship between disease severity and variability was weaker during music (slope = 0.24, SE = 0.19, 95% CI [−0.14, 0.62]).than silence (slope = 0.56, SE = 0.19, 95% CI [0.18, 0.93]). This difference was significant (Silent − Music = 0.32, SE = 0.11, p = .004) (Figure 4. C).

### Stride Length Variability

For stride length variability, there was a significant main effect of cue (F(1,712) = 109.17, p < .001, R²β = .13), with greater variability during music than silence (post-hoc: p < .0001). (Figure 2.B). No other main effects were significant (all p > .12). A significant Cue × Rhythm interaction (F(1,712) = 7.95, p = .005, R²β = .01) and a Cue × Group × Rhythm interaction (F(1,712) = 6.99, p = .008, R²β = .01) were observed. Post-hoc analysis showed that among older adults, higher rhythmic ability predicted greater stride length variability during silent walking (slope = 0.16, SE = 0.28, 95% CI [−0.40, 0.73]), whereas during music walking the relationship was negative (slope = −0.49, SE = 0.28, 95% CI [−1.05, 0.07]). The difference between conditions was significant (Silent Older − Music Older = 0.65, SE = 0.17, p = .001), indicating that music reversed the direction of the rhythmic ability variability relationship in older adults, and no significant effects were seen for the PD group (Figure 3. H). No significant differences were observed across any other pairwise comparisons between groups or cue conditions (all p > .47). In the PD only model, greater disease severity was associated with higher stride length variability (F(1,32.93) = 5.42, p = .026, R²β = .141).

### Stability variables Stride Width

Stride width showed a significant main effect of time (F(2,712) = 4.11, p = .02, R²β = .01), with stride width being wider immediately post-stimulation compared to during stimulation (p = .02), while the 15-minute post-stimulation timepoint did not significantly differ from either (both ps > .32) (Figure 1. D). There were no significant main effects of stimulation type, cue, group, rhythm, or sex (all ps > .06). The only significant interaction was Stimulation × Group (F(1,712) = 8.16, p = .004, R²β = .01), indicating that the anodal versus sham difference in stride width differed between groups (sham – anodal, Older – PD; p = .006); post-hoc analysis showed that older adults showed a small increase in stride width under anodal relative to sham stimulation, whereas individuals with PD showed the opposite pattern, with narrower stride width under anodal compared to sham, with a larger sham to anodal change in the PD group relative to older adults.

### Double Support Time

Double support time showed significant main effects of stimulation, cue, time, rhythm, and sex. Double support time was shorter during anodal than sham stimulation (F(1,712) = 9.88, p = .002, R²β = .01), and shorter during music than silence (F(1,712) = 44.68, p < .001, R²β = .06). For time (F(2,712) = 29.32, p < .001, R²β = .08), double support time was also shorter at 15 minutes post-stimulation compared to both during stimulation (p < .0001) and immediately post-stimulation (p < .0001), with no difference between the latter two (p = .46) (Figure 1. E). Participants with stronger rhythmic abilities showed shorter double support times (F(1,65) = 5.59, p = .02, R²β = .08), and relative to baseline, females showed shorter double support times than males (F(1,65) = 5.98, p = .02, R²β = .08). There was no significant main effect of group (F(1,65) = 0.08, p = .78).

These effects were qualified by significant Cue × Time (F(2,712) = 12.17, p < .001, R²β = .03), Stimulation × Rhythm (F(1,712) = 4.26, p = .04, R²β = .01), Cue × Rhythm (F(1,712) = 11.79, p < .001, R²β = .02), Stimulation × Group × Rhythm (F(1,712) = 4.39, p = .04, R²β = .01), and Stimulation × Cue × Group × Rhythm (F(1,712) = 4.51, p = .03, R²β = .01) interactions, the latter being the highest order significant effect. Post-hoc analysis for the Cue × Time interaction showed that the music-silence difference in double support time was larger at 15 minutes post-stimulation compared to immediately post-stimulation (p < .0001) and during stimulation (p = .05), with a significant difference also between stimulation and post-stimulation (p = .006). For the Stimulation × Cue × Group × Rhythm interaction, post-hoc analysis showed that the rhythm slope was most negative under anodal music conditions in the PD group (slope = −2.72, SE = 1.01, 95% CI [−4.71, −0.73]) compared to the sham silent condition in PD (slope = 0.23, SE = 1.01, 95% CI [−1.76, 2.23]). The only significant pairwise contrast after correction was between sham-silent PD and anodal-music PD (difference = 2.95, SE = 0.73, p = .001), indicating that anodal stimulation combined with music strengthened the negative relationship between rhythmic ability and double support time in PD (Figure 3.F). All other comparisons were non-significant (all p > .11).

### Subjective experience of walking to the beat and tDCS feeling

Participants’ subjective ratings of difficulty in synchronizing to the beat when walking to music and sensation of stimulation intensity were compared between the anodal and sham sessions. For difficulty synchronizing, the mean rating was 41.9 in the anodal session and 37.2 in the sham session. A Welch two-sample t-test indicated no significant difference between sessions (t(63.89) = 0.59, p = 0.56, 95% CI [-11.18, 20.51]). Similarly, for the tDCS sensation intensity, the mean rating was 39.9 in the anodal session and 33.4 in the sham session, with no significant difference observed (t(35.98) = 0.97, p = 0.34, 95% CI [-7.07, 19.92]). These results suggest that participants’ subjective perceptions of task difficulty and stimulation intensity were comparable between anodal and sham conditions.

## Discussion

Here we investigated whether anodal tDCS over the SMA enhances the effects of rhythmic auditory cueing in individuals with Parkinson’s disease (PD) and healthy older adults. We also tested whether these effects differed during online stimulation versus after stimulation ceased, as well as the influence of individual differences in rhythmic ability and disease severity. By combining tDCS stimulation with music-based cueing, we aimed to disentangle whether these two interventions for improving gait would show additive or interactive effects. Overall, the findings provide three main insights. First, rhythmic auditory cueing improved temporal aspects of gait, with particularly strong effects on cadence, stride velocity, and double support time, especially 15 minutes-post stimulation. Second, anodal SMA stimulation selectively improved gait stability and temporal variability, particularly reducing stride time variability and stride width, with effects that were most pronounced in individuals with PD, reducing differences relative to healthy older adults during sham stimulation. Third, rhythmic ability and disease severity modulated responsiveness to rhythmic auditory cueing and tDCS.

Music cueing affected temporal gait parameters, increasing cadence and reducing double support time, with the largest effects observed 15 minutes post-stimulation. However, these temporal gains were accompanied by shorter stride lengths and increased variability in spatial gait measures, indicating a potential spatiotemporal trade-off where faster step timing may come at the cost of increased spatial variability. In contrast, anodal SMA-tDCS reduced gait variability, including stride time and stride length variability, as well as overall stride width in people with PD, particularly in individuals with greater disease severity. These reductions in variability were present during stimulation, and, for stride time variability, persisted 15 minutes after stimulation had ended. Importantly, both cueing- and stimulation effects were moderated by individual differences: participants with stronger rhythmic abilities had larger music-related increases in cadence, whereas disease severity worsened spatial gait variability during music-cued walking. Greater disease severity was also associated with larger reductions in stride time and stride length variability following anodal tDCS. While rhythmic cueing affected gait timing in both groups, individuals with PD (who overall showed slower and more variable gait than controls) showed larger reductions in variability with SMA-tDCS. Together, these findings suggest that rhythmic cueing primarily influenced temporal gait parameters (cadence and double support time), whereas SMA-tDCS primarily influenced gait variability measures, indicating that the two interventions affect different aspects of gait control.

### Spatiotemporal outcomes

Music produced changes in spatiotemporal gait parameters, increasing cadence and reducing stride length and double support time. These findings align with previous evidence that rhythmic auditory cueing improves temporal aspects of gait in PD by providing an external pacing signal (Del Olmo & Cudeiro, 2005; Howe et al., 2003; Lee & Ko, 2023; Spaulding et al., 2013). Past studies have shown that faster rhythmic cues can increase gait velocity, cadence and stride length (Ghai et al., 2018; Harrison et al., 2025; McIntosh et al., 1997; Thaut et al., 1996). Longer-term music interventions have similarly been shown to sustain improvements in stride time, cadence, and motor symptom severity (De Bruin et al., 2010), and meta-analytic work further corroborates that rhythmic cueing improves walking velocity, stride length, and cadence across diverse cueing paradigms (Lee & Ko, 2023).

However, the effects of rhythmic cueing on individual gait parameters are not uniform across studies or populations. In stroke, tempo manipulations of 10% and 20% faster than preferred cadence have been shown to significantly improve gait velocity and cadence, while slower tempi reduced them, though without a parallel increase in stride length at the faster tempi (Cha et al., 2014). In PD, auditory cueing has been linked to improved velocity and stride length but not cadence (Ghai et al., 2018; Wang et al., 2022). In contrast, a study showed that 115% cadence cues increased gait speed and cadence without altering step length (Hoppe et al., 2020). Differently from prior work, here music significantly decreased stride length, possibly reflecting a compensatory strategy to maintain rhythmic entrainment and temporal precision. This is also seen in early PD, when participants develop compensatory mechanisms by increasing cadence and decreasing swing time (Panyakaew & Bhidayasiri, 2013).

Anodal tDCS over the SMA in our single-session design produced modest but reliable effects on gait timing, increasing cadence relative to sham, while effects on gait velocity and spatial parameters were less consistent. These timing effects may align with the SMA’s role in internally generated movement sequencing and temporal control (Cona & Semenza, 2017; Cunnington et al., 1996; Goldberg, 1985) and may explain why stimulation influenced cadence rather than stride length or velocity. The magnitude of the tDCS effects was smaller than that induced by rhythmic cueing, a pattern consistent with previous studies reporting mixed or selective effects of tDCS on gait when applied in isolation (Benninger et al., 2010). However, our results are consistent with prior work that pairs tDCS with physical or cue-based training and finds that stimulation effects can amplify behavioural intervention effects and persist (Costa-Ribeiro et al., 2016; Kaski et al., 2014), suggesting that tDCS may serve as a neuromodulatory facilitator of gait timing, rather than as an independent driver of gait acceleration.

Notably, the strongest cueing effects on cadence and stride velocity were 15 minutes post-stimulation. Because these delayed effects were not modulated by stimulation type, they are unlikely to reflect tDCS aftereffects. The delay may have provided a recovery time after the attentional and physical demands of walking during stimulation. However, these effects were quantified as changes relative to an initial baseline, collected when participants were presumably well-rested. Therefore, they may reflect post-exertional arousal or a carryover effect of rhythmic cueing, consistent with other work finding persistent benefits after stimulation (Thaut et al., 1996).

Individual rhythmic ability also influenced cueing responses. Participants with better rhythmic skills showed greater cadence increases during music, consistent with evidence that beat perception and synchronization abilities predict responsiveness to rhythmic auditory stimulation (Bella et al., 2017, 2018; Cochen De Cock et al., 2018). These abilities likely reflect the efficiency of auditory-motor coupling mechanisms, including beat extraction, temporal prediction, and error correction, that support movement alignment to external rhythms (Grahn & Rowe, 2009; Zatorre et al., 2007). In PD, basal ganglia dysfunction disrupts internal timing (Breska & Ivry, 2018; Freeman et al., 1993; Nombela et al., 2013), which means that individuals with stronger externally driven timing abilities may be better positioned to use rhythmic cues in a compensatory way, suggesting that the efficacy of rhythmic auditory cueing depends, at least in part, on individual auditory-motor capacities.

### Variability outcomes

A key finding was that tDCS reduced variability of stride time and length. Increased gait variability is a strong predictor of fall risk in PD (Hausdorff, 2009; Hausdorff et al., 2001; Toebes et al., 2012), making these effects clinically relevant even when modest. Variability reductions were most evident during stimulation, and, for stride time variability, persisted at 15 minutes post-stimulation, suggesting a stabilizing influence of SMA stimulation on gait. This finding is consistent with emerging, though mixed, evidence that neuromodulation can influence gait regularity. For instance, anodal tDCS over the prefrontal cortex combined with aerobic exercise has been shown to decrease step time variability (Conceição et al., 2021), and a recent systematic review identified improvements in dual-task gait performance following dorsolateral prefrontal tDCS in several studies (Moeyersons et al., 2025). However, other reviews found no consistent or clinically significant benefits when tDCS was added to gait training (Nascimento et al., 2021; Pol et al., 2021), suggesting that effects on variability may depend on individual responsiveness and stimulation parameters.

Causal evidence supports a role of the SMA in the temporal and attentional control of gait, particularly in planning and initiation processes (Goh et al., 2019; Richard et al., 2017). In healthy adults, inhibitory stimulation of the SMA disrupts anticipatory postural adjustments and step timing, highlighting its contribution to gait planning rather than execution alone (Richard et al., 2017). Facilitatory stimulation over the SMA has also been shown to improve dual-task walking performance, whereas stimulation of other regions such as the dorsolateral prefrontal cortex or primary motor cortex does not yield similar benefits (Goh et al., 2019). In clinical populations, evidence is more variable but suggests that effects depend on task context and stimulation protocol. In PD, single session anodal tDCS over the SMA in isolation has shown limited effects on gait (Kaski et al., 2014), whereas combining SMA stimulation with gait or cueing interventions across multiple sessions has been associated with improvements in gait velocity, balance, and functional mobility, as well as prolonged retention of training effects (Costa-Ribeiro et al., 2017; Kaski et al., 2014; Lee & Kim, 2021). Similarly, repetitive TMS over the SMA has been reported to reduce freezing of gait and improve gait outcomes, although findings are not entirely consistent (Kim et al., 2018; Lu et al., 2018; Mi et al., 2019).

In contrast, music cueing increased variability in stride length and velocity, especially in individuals with poorer rhythmic abilities or greater disease severity. This pattern is consistent with previous findings that while rhythmic cueing often enhances speed and cadence, it can also introduce instability in stride timing—particularly among individuals with impaired beat perception (Bella et al., 2017; Dotov et al., 2017).

Indeed, responses to rhythmic auditory stimulation are variable: some PD patients show significant reductions in stride-to-stride variability (Del Olmo & Cudeiro, 2005; Hausdorff et al., 2007), whereas others display minimal or even adverse effects (Harrison & Earhart, 2023). The variability in outcomes likely reflects individual differences in the capacity for sensorimotor entrainment, which is shaped by both musical experience (Bella et al., 2018; Cochen De Cock et al., 2018) and the heterogeneity within the disease itself, including differences in dopaminergic loss, cognitive profile, and stage of disease progression.

### Stability outcomes

Stability measures provided additional insights into the effects of rhythmic cueing and tDCS on gait control. Double support time decreased with music, particularly 15 minutes post-stimulation, likely reflecting faster step-to-step transitions associated with faster cadence. One study reported shorter double support time after cueing, interpreting it as greater confidence in single-limb support and improved weight transfer, with its effects persisting beyond the immediate cueing period (Bryant et al., 2009). However, the relationship between double support time and fall risk is complex. In PD, longer and more variable double support time is associated with instability and falls (Callisaya et al., 2011; Cole et al., 2010), but absolute reductions may also reflect increased walking speed rather than enhanced postural control. Thus, the observed reduction in double support time under music may well be a consequence of faster cadence, rather than a direct indicator of improved balance.

Stride width, by contrast, was selectively modulated by SMA-tDCS. Anodal stimulation resulted in narrower width compared to sham, independent of cueing, time point, group, rhythmic abilities, or disease severity. Because stride width is commonly interpreted as a marker of mediolateral balance control and cautious gait strategies (Roden-Reynolds et al., 2015), this finding suggests that SMA stimulation may influence aspects of gait stability that are less sensitive to rhythmic timing cues. The absence of cueing effects on stride width aligns with prior reports showing inconsistent or negligible effects of rhythmic auditory cueing on stride width (Wang et al., 2022), likely because such control relies more heavily on postural and proprioceptive mechanisms than on temporal gait regulation (Bonnet, 2014; Roden-Reynolds et al., 2015).

Individual rhythmic ability further modulated double support time, with participants showing stronger rhythmic skills exhibiting larger reductions during both music and anodal stimulation. These findings reinforce the view that rhythmic auditory cueing and SMA-tDCS act through partially distinct mechanisms: music affects temporal coordination and step timing, while tDCS modulates gait stability, particularly stride width.

### The SMA in gait and cueing

Although the present study was not designed to directly probe neural mechanisms, the pattern of behavioral effects is consistent with a role for the SMA as a stabilizing node in gait control. The SMA is critically involved in internally timed movement sequences, gait initiation, and rhythm perception (Cannon & Patel, 2021; Grahn & Brett, 2007; Jahanshahi et al., 1996), and its activity is disrupted in PD (Muthukrishnan et al., 2019; Sabatini et al., 1998). Here, anodal tDCS over the SMA influenced cadence, stride time, variability, and stride width, suggesting an effect on the consistency and reliability of gait execution rather than on overall gait speed. Specifically, these findings are consistent with a role for the SMA in reducing temporal and spatial variability in gait execution.

In parallel, rhythmic auditory cueing provided an external temporal scaffold that increased pacing but also introduced variability in spatial parameters. These complementary effects support a model in which rhythmic cueing supplies an external timing signal that compensates for impaired basal ganglia-SMA circuits (Nombela et al., 2013), while SMA stimulation enhances the stability of internally generated timing and coordination. From this perspective, tDCS does not amplify cueing effects directly, but may increase the reliability of these internal timing signals, reducing gait variability.

### Group differences: healthy older adults versus Parkinson’s Disease

Comparisons between healthy older adults and individuals with PD revealed both shared and disease-specific responses to auditory cueing and SMA-tDCS. Rhythmic cueing increased cadence in both groups, confirming preserved auditory-motor entrainment across aging and PD. However, individuals with PD showed a reduced ability to increase cadence relative to healthy older adults, particularly during music, consistent with impaired motor function and temporal flexibility associated with basal ganglia network dysfunction (Hausdorff, 2009; Morris et al., 1996; Sabatini et al., 1998).

In contrast, stimulation effects on stability measures were specific to the PD group. Under sham stimulation, individuals with PD exhibited wider steps and longer double support time than controls, reflecting compensatory strategies for postural instability (Bloem et al., 2004; Curtze et al., 2016; Morris et al., 2001). Anodal SMA-tDCS reduced stride width and double support time in the PD group, whereas healthy older adults showed little change. Similarly, reductions in gait variability measures were primarily observed in PD, supporting the idea that SMA stimulation may attenuate pathological gait features rather than enhance already efficient control in healthy aging.

### Clinical implications

These findings suggest that rhythmic auditory cueing and SMA-tDCS may serve complementary roles in gait rehabilitation for PD. Music cueing appears to enhance gait pacing, whereas SMA stimulation selectively influences gait stability and variability. Thus, a combination of both may be more effective than either alone. These approaches may complement pharmacological and surgical treatments, which often have limited efficacy for gait disturbances in PD (Bella et al., 2015; Grabli et al., 2012).

## Limitations and future directions

Several limitations should be noted. This was a single-session study, limiting conclusions about long-term effects; prior research indicates cumulative effects of repeated active tDCS sessions (Pol et al., 2021; Rostami et al., 2020), which we did not perform. Participants were tested in the ON medication state, which may interact with stimulation and music effects (but is also the state in which most interventions would be typically delivered). Music tempo was fixed at 10% faster than baseline cadence, but future work should explore individualized tempo and musical preference. Moreover, no direct neural measures, such as fMRI, were included to verify SMA modulation. Thus, future studies could investigate multi-session interventions, ON/OFF-medication testing, more individualized musical parameters, and direct neural measures to better characterize mechanisms and optimize intervention protocols.

## Conclusion

In summary, rhythmic auditory cueing with music enhances temporal aspects of gait in PD, particularly increasing cadence, whereas anodal tDCS over the SMA also increased cadence, but selectively reduced gait variability and narrowed stride width. These effects were additive rather than interactive, revealing a dissociation between gait pacing and gait stability. Together, these findings highlight the importance of targeting distinct components of gait control and support the potential of innovative, multi-modal rehabilitation approaches for gait rehabilitation in PD.

## Data Availability

All data produced in the present study are available upon reasonable request to the authors.

## Supplementary information

**Table 1.** Means and SDs by walking variable for PD.

| Walking Variable | Walk Time | Sham_Silent Mean(SD) | Sham_Music Mean(SD) | Anodal_Silent Mean(SD) | Anodal_Music Mean(SD) |
| --- | --- | --- | --- | --- | --- |
|  |  | PD | PD | PD | PD |
| **Cadence** | Stimulation | -0.02 (3.64) | 4.4 (4.73) | 1.26 (2.42) | 5.18 (4.69) |
|  | Post-Stim | 0.59 (3.93) | 3.84 (5.79) | 1.88 (3.13) | 4.63 (5.85) |
|  | 15-min Post-Stim | 2.01 (4.17) | 7.13 (4.25) | 2.71 (3.12) | 7.55 (4.23) |
| **Stride Length** | Stimulation | -0.3 (4.81) | -6.65 (10.35) | 0.07 (4.21) | -6.35 (11.01) |
|  | Post-Stim | -0.24 (6.19) | -5.86 (12.12) | -0.53 (6.13) | -6.49 (9.78) |
|  | 15-min Post-Stim | 0.81 (4.89) | -0.76 (12.94) | -0.1 (6.82) | -3.45 (11.98) |
| **Stride Velocity** | Stimulation | -0.12 (7.53) | -2.67 (11.22) | 1.3 (5.13) | -1.74 (11.72) |
|  | Post-Stim | 0.5 (8.44) | -2.53 (12.51) | 1.47 (7.55) | -2.49 (10.04) |
|  | 15-min Post-Stim | 3.19 (7.79) | 6.4 (14.74) | 2.57 (8.29) | 3.62 (13.66) |
| **COV Stride Time** | Stimulation | 2.88 (1.44) | 3.42 (1.57) | 2.41 (1.18) | 2.96 (0.96) |
|  | Post-Stim | 3.09 (3.37) | 3.65 (3) | 2.85 (3.1) | 2.93 (1.26) |
|  | 15-min Post-Stim | 2.77 (2.65) | 2.43 (0.99) | 2.41 (1.29) | 2.69 (0.87) |
| **COV Stride Length** | Stimulation | 3.92 (1.67) | 5.53 (2.94) | 3.36 (1.4) | 5.29 (3.14) |
|  | Post-Stim | 4.13 (3.18) | 5.54 (4.32) | 3.79 (3.13) | 4.77 (3.14) |
|  | 15-min Post-Stim | 3.86 (3.14) | 4.97 (2.57) | 3.55 (1.64) | 4.96 (2.47) |
| **COV Stride Velocity** | Stimulation | 4.47 (1.35) | 5.59 (2.6) | 4.18 (1.36) | 5.51 (3.05) |
|  | Post-Stim | 4.44 (1.95) | 5.13 (2.6) | 4.29 (1.62) | 4.81 (2.93) |
|  | 15-min Post-Stim | 4.07 (1.73) | 5.17 (2.62) | 4.46 (1.86) | 5 (2.31) |
| **Stride Width** | Stimulation | 11.04 (48.86) | 16.93 (42) | 0.4 (24.74) | 10.65 (30.3) |
|  | Post-Stim | 13.29 (44.12) | 24.13 (44.69) | 13.86 (24.72) | 13.93 (35.14) |
|  | 15-min Post-Stim | 13.44 (55.96) | 18.45 (52.85) | 7.03 (28.38) | 6.55 (28.85) |
| **Double Support Time** | Stimulation | 2.56 (7.01) | -0.32 (10.76) | 1.1 (5.25) | -1.15 (10.51) |
|  | Post-Stim | 0.67 (7.14) | -0.46 (11.19) | -0.15 (7.17) | -0.14 (11.69) |
|  | 15-min Post-Stim | -1.1 (7.58) | -7.53 (12.24) | -0.79 (8.12) | -6.83 (13.32) |

**Table 2.**
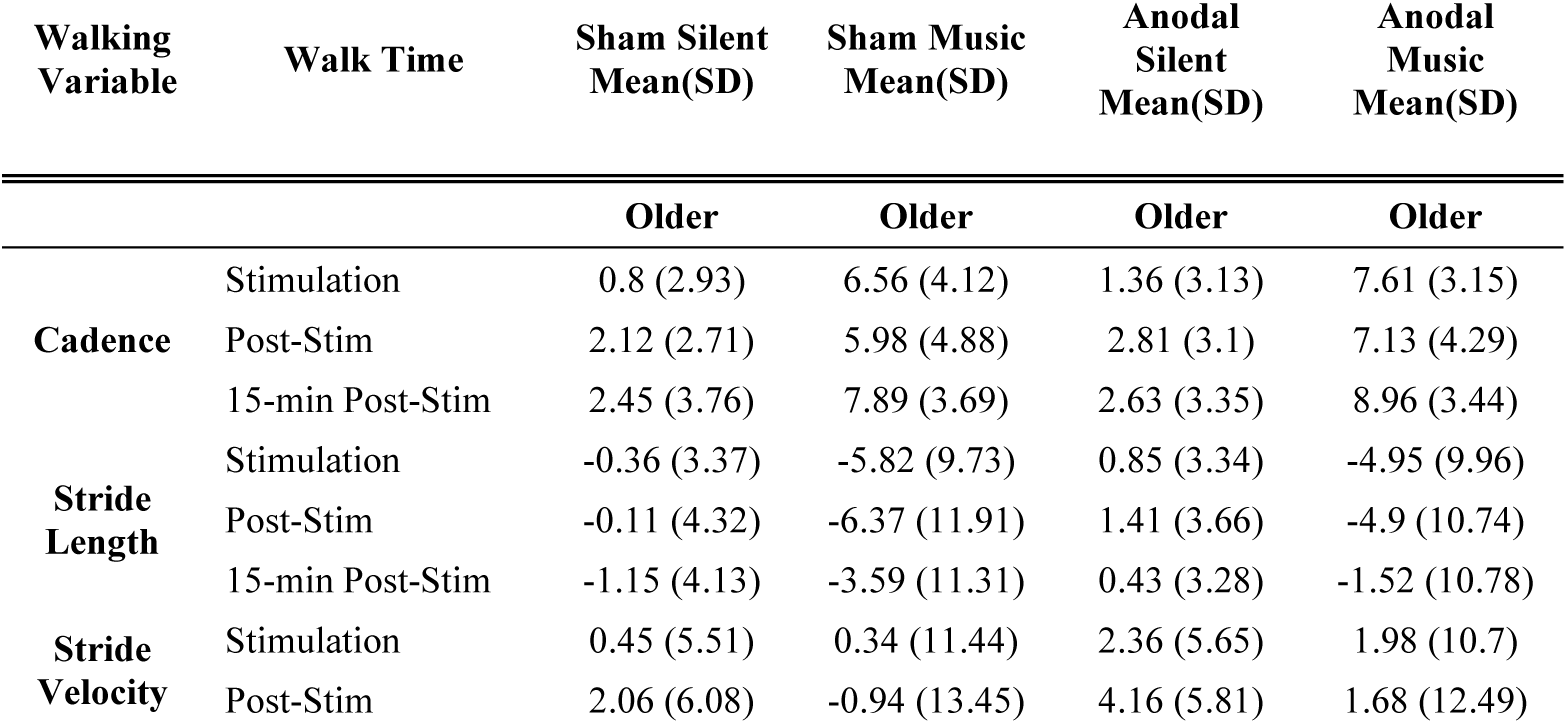
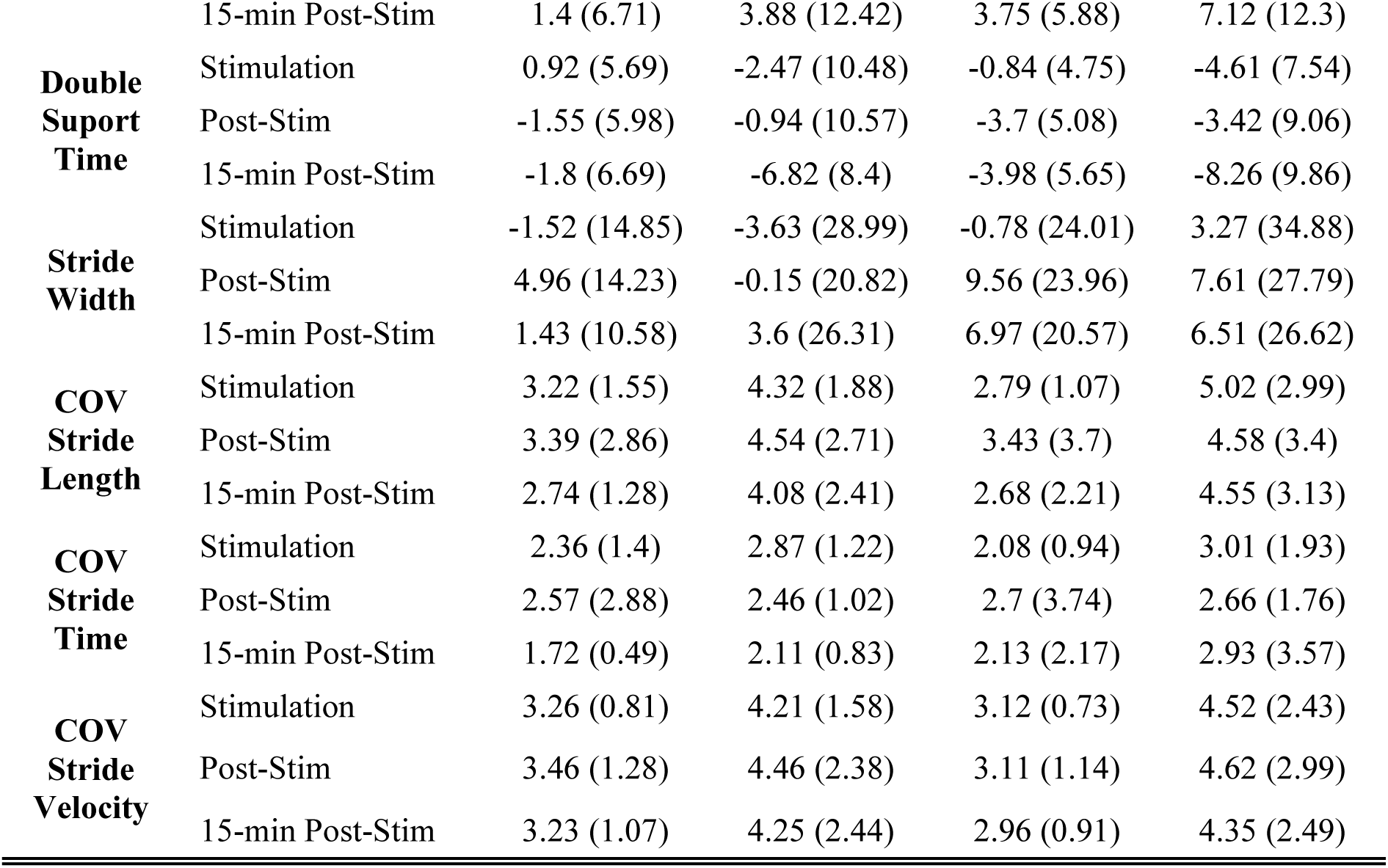
Means and SDs by walking variable for older adults.

## Notes

### Competing Interest Statement

The authors have declared no competing interest.

### Clinical Trial

The study was designed as a mechanistic investigation of neuromodulation and auditory-motor coupling rather than a clinical efficacy trial. This study is a prospective interventional experiment using a within-subject crossover design to investigate the effects of tDCS and auditory cueing on gait in Parkinson's disease. It is not a clinical trial aimed at evaluating a therapeutic intervention for clinical use and therefore was not registered in a clinical trials registry.

### Funding Statement

This work was supported by the Natural Sciences and Engineering Research Council of Canada (NSERC) and by the Parkinson Society Southwestern Ontario-Mitacs Graduate Student Scholarship.

### Author Declarations

The Western University Health Science Research Ethics Board (HSREB) has reviewed and approved this work. Project ID 123250.

### Summary of Updates

- Figures have been updated to include both the control and Parkinson's disease (PD) groups. - The Results section has been streamlined and now focuses solely on comparisons between the two groups. - The Discussion has been revised to better reflect and interpret the updated results.

